# Combined oropharyngeal/nares and nasopharyngeal swab sampling remain effective for molecular detection of SARS-CoV-2 Omicron variant

**DOI:** 10.1101/2022.02.13.22270891

**Authors:** Glenn Patriquin, Jason J. LeBlanc, Holly A. Gillis, Gregory R. McCracken, Janice J. Pettipas, Todd F. Hatchette

## Abstract

The world has experienced several waves of SARS-CoV-2 variants of concern (VoCs) throughout the COVID-19 pandemic since the first cases in December 2019. The Omicron VoC has increased transmission, compared to its predecessors, and can present with sore throat and other cold-like symptoms. Given the predominance of throat symptoms, and previous work demonstrating better sensitivity using antigen-based rapid detection tests when a throat swab is included in the standard nasal sampling, this quality improvement project sought to ensure ongoing suitability of both combined oropharyngeal/nares (OPN) and nasopharyngeal (NP) swab sampling used throughout the pandemic. Consenting participants meeting Public Health testing criteria (mostly symptomatic or a close contact of a known case) were enrolled, and paired NP and OPN swabs collections were subjected to nucleic acid amplification testing (NAAT). Comparing paired specimens from 392 participants sensitivity of NP swabs was 89.1% (95% CI, 78.8-94.9), and that of OPN was 98.4% (95% CI: 90.9->99.9) (p-value 0.052). This project demonstrated that both NP and combined OPN swabs detected the Omicron variant with similar sensitivity by NAAT, supporting the continued use of either swab collection for SARS-CoV-2 molecular detection.

## Short communication

Since COVID-19 was first described in December 2019, several SARS-CoV-2 variants of concern (VoCs) have been detected, each with unique properties, both in terms of symptomatology and transmissibility. Prior to the emergence of VoCs, RT-PCR of testing nasopharyngeal (NP) swabs was the gold standard approach to testing in community and hospitalized patients, with the addition of lower tract specimens to increase sensitivity in those with pulmonary disease^1^. Early in the global response, the reduced availability of NP swabs necessitated the validation of several combined oropharyngeal/nares (OPN) swabs as an alternative sampling method, which demonstrated acceptable performance when compared to the NP^2,3^. Recently the Omicron VoC (lineage B.1.1.529)^4^ has become predominant in many regions in the world and is notable for its increased transmissibility, reduced vaccine effectiveness^5,6^, and possible differences in tissue tropism^7–9^. The propensity to cause sore throat and cold-like symptoms^10^, along with previous work showing improved sensitivity with inclusion of a throat sample in combination with nasal sampling for use with antigen-based rapid testing devices (Ag-RDTs)^11,12^, prompted this quality improvement initiative.

Community members who met eligibility criteria for molecular diagnostic testing by Public Health (Table S1) were asked to participate in this project. Participants were adults recruited from one of two testing streams during a four-day period in January of 2022, in Nova Scotia, Canada. Enrollment was either facilitated by Public Health through the Public Health Mobile Testing Unit deployed for outbreak investigations in multiple communities, or through urban-based COVID-19 testing centres. In both streams, participants had NP sampling for nucleic acid amplification testing (NAAT), using a standardized sampling technique with a flocked swab (https://vimeo.com/516853275/c67017fd3a), which was placed in viral transport media (VTM) (Copan Diagnostics, Inc., (Murrietta, CA), or AccuViral Collection Kit, (AccuGene, San Diego, CA)). Following verbal consent, an additional combined OPN swab was collected as previously described^2,3,13^, using a CLASSIQSwab fiber-tip swab from Copan Italia (Brescia, Italy), placed into the same type of VTM.

NP and OPN samples were immediately transported to a central laboratory and refrigerated at 4° Celsius, where NAAT was performed within 12h according to manufacturer instructions. Initially, both NP and OPN samples were tested using the Aptima SARS-CoV-2 Assay on the Panther System (Hologic Inc., San Diego, CA). NP and OPN swabs with negative Panther results were confirmed as negative with the SARS-CoV-2 Test on the Cobas 6800 instrument (Roche Diagnostics, Rotkreuz, Switzerland). NP or OPN swabs with positive results, as well as the paired specimen regardless of its result, were subsequently tested using both the Roche 6800 assay and the Xpert Xpress SARS-CoV-2 test (Cepheid, Sunnyvale, CA). Swab result was compared to a consensus reference standard derived from all NAAT test results. For each swab, a positive result was defined as SARS-CoV-2 detection in at least two NAATs (Table S2). To categorize swab results as true positive, true negative, false positive, and false negative results (to determine sensitivity and specificity), each swab was compared to the case result, established at the level of the individual. McNemar’s test with Yate’s correction (https://www.omnicalculator.com/) was used to determine statistical significance between NP and OPN swab collections. This project was deemed a quality improvement initiative and was exempt from Research Ethics Board review (file number 1027644).

Overall, 392 participants with paired NP and OPN samples were analyzed, with concurrent results between swab types seen for the majority of individuals (Table). The overall positivity rate was 16.3%, with 328 individuals negative by both swab types, and 56 with positive results from both swab types. Eight people had conflicting results, with one SARS-COV-2 detection identified by NP alone, and seven detected by OPN alone. Sensitivity and specificity were 89.1% (95% confidence interval (CI): 78.8-94.9%) and 100% (95% CI: 92.5-100), and 98.4% (95% CI: 90.9->99.9%) and 100% (95% CI: 93.1-100), for NP and OPN swabs, respectively. However, this difference between the swab types was not statistically significant (p-value, 0.052). All OPN samples with Ct values <32 were submitted for next-generation sequencing at the National Microbiology Laboratory (NML) (Winnipeg, Manitoba). Of 59 samples submitted, four sequencing reactions failed, and the remaining 55 samples were characterized as the VoC, Omicron (50 of the BA.1 lineage, and five of the BA.1.1 lineage).

**Table.** SARS-CoV-2 nucleic acid amplification test results of paired nasopharyngeal and oropharyngeal/nares swabs.

|  | Number of participants |
| --- | --- |
| Positive by NP and OPN | 56 |
| Positive by NP only | 1 |
| Positive by OPN only | 7 |
| Negative by NP and OPN | 328 |
| *p-value* (NP vs. OPN)* | 0.052 |

\*McNemar's test with Yate's correction; NP, nasopharyngeal; OPN, oropharyngeal/nares.

The COVID-19 pandemic has been fraught with various challenges; each wave uniquely testing our ability to detect, contain, and prevent infection. With Omicron, questions have been raised regarding altered tissue tropism, which caused people to question the optimal anatomical location for SARS-CoV-2 detection. In this quality project, there was no significant difference between NP or OPN specimens for the detection of SARS-CoV-2 when using NAAT methods.

This conclusion differs from our recent study in which the sensitivity of Ag-RDTs for detecting SARS-CoV-2 was higher when sampling the throat and nose (versus nasal sampling alone). However, the results of that study are not directly comparable to this population because the current project compared the nasopharynx and the oropharynx/nares, whereas the Ag-RDT study compared bilateral nares swabbing with an OPN swab. It may be that the anatomical proximity of the nasopharynx to the oropharynx rendered a difference between sampling these sites insignificant. In addition, perhaps NAAT testing, being more sensitive than Ag-RDT testing was not subject to noticeable differences between the sites. In this project, although not significant, there were some cases detected by OPN that were missed by NP sampling. It is possible that some of these results reflect the difficulty in performing an NP swab consistently, as compared to the relative ease of performing an OPN swab; however, given a lack of a reliable molecular host target for sample quality, the possibility of discrepant results based on differences in anatomical sites is also possible^7–9^.

This project was limited in several ways. As it was a quality initiative, clinical information for participants was not available, therefore symptomatology, timing of swab collection from recent exposures, and the impact of co-morbid conditions or immunization status could not be established. Also, despite policies and training in the standardization of performing NP swabs, the quality of the technique could not be directly assessed.

Ultimately, despite evidence indicating that combined sampling of the throat and nares can significantly increase SARS-CoV-2 detection by Ag-RDTs compared to nasals swabs, the performance characteristics of OPN swabs did not differ from the reference NP sampling method for NAAT detection of SARS-CoV-2. Importantly, this work demonstrates the ongoing suitability of either NP or OPN swab for molecular detection of SARS-CoV-2, even if Omicron potentially has altered tissue tropism.

## Data Availability

All data produced in the present work are contained in the manuscript

## Acknowledgements

The authors thank the constant efforts of those working in the COVID-19 Testing Centres and the Public Health Mobile Units for their support in this project. Your dedication and enthusiasm for projects such as these are instrumental for the ongoing care of the community. The authors would also like to thank front line staff in the Division of Microbiology, Department of Pathology and Laboratory Medicine, Nova Scotia Health, for their support for this project and unwavering efforts throughout the pandemic. Thank you to the National Microbiology Laboratory for providing surveillance sequencing data.

---

**Table S1.**
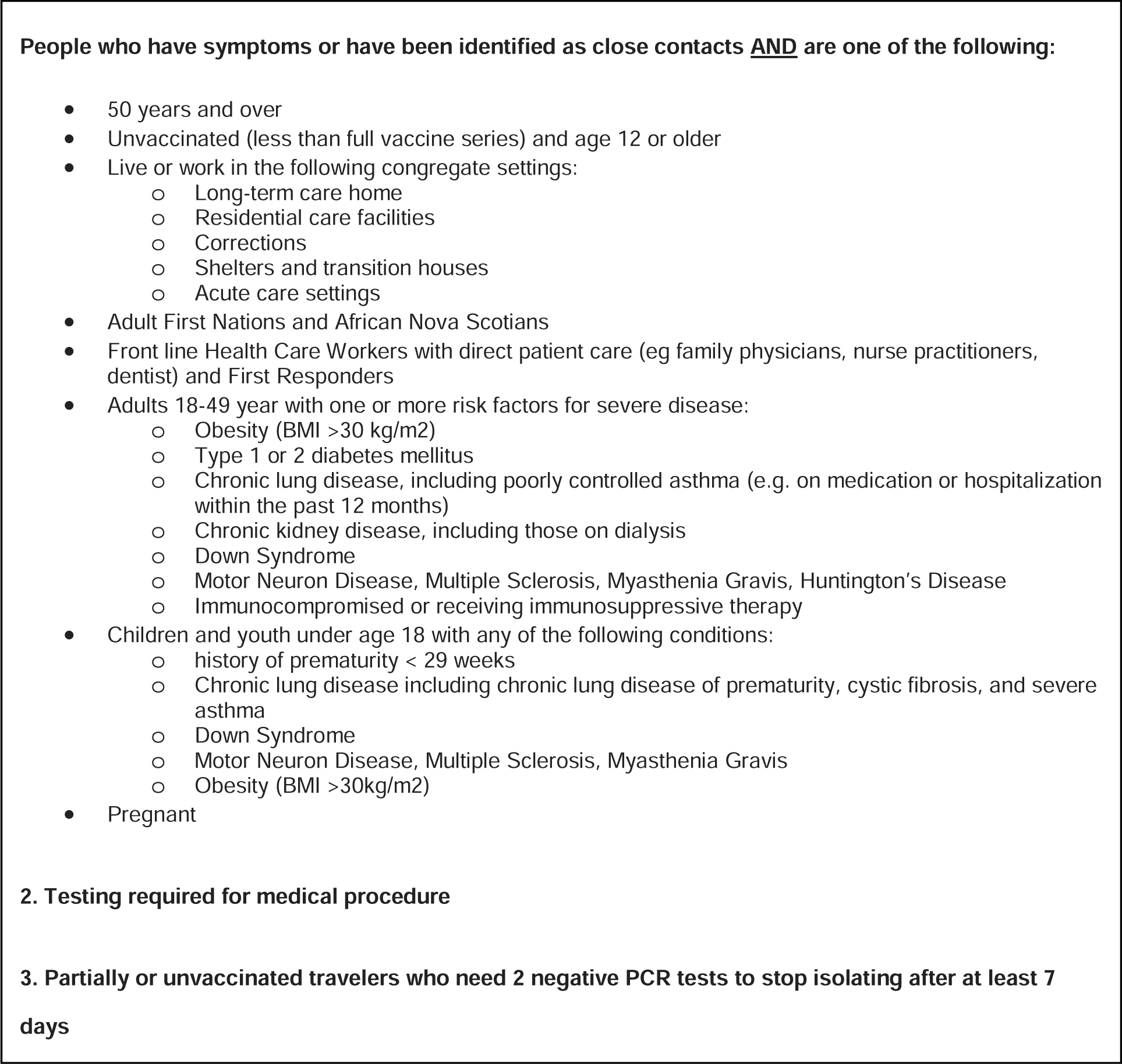
Provincial Public Health eligibility criteria for COVID-19 testing by polymerase chain reaction, at the time of data collection.

**Table S2.** Discrepant results between NP and OPN swab collections.

| Sample | NP swab |  |  |  |  |  |  |  |  | OPN swab |  |  |  |  |  |  |  |  |
| --- | --- | --- | --- | --- | --- | --- | --- | --- | --- | --- | --- | --- | --- | --- | --- | --- | --- | --- |
|  | Panther |  | 6800 |  |  | Xpert |  |  | Overall Result | Panther |  | 6800 |  |  | Xpert |  |  | Overall Result |
|  | RLU | Result | Ct Orf1a/b | Ct E gene | Result | Ct E gene | Ct N2 gene | Result |  | RLU | Result | Ct Orf1a/b | Ct E gene | Result | Ct E gene | Ct N2 gene | Result |  |
| 1 | 906 | POS | 41.6 | 42.7 | POS | 0 | 0 | NEG | POS | NEG | NEG | 0 | 0 | NEG | 0 | 0 | NEG | NEG |
| 2 | NEG | NEG | 0 | 0 | NEG | 0 | 37.87 | POS | NEG | 812 | POS | 0 | 41.7 | POS | 0 | 37.25 | POS | POS |
| 3 | NEG | NEG | 39 | 41.8 | POS | 0 | 0 | NEG | NEG | 1265 | POS | 35.1 | 38.3 | POS | 35.76 | 37.1 | POS | POS |
| 4 | NEG | NEG | 0 | 0 | NEG | 0 | 0 | NEG | NEG | 1227 | POS | 38.6 | 40.7 | POS | 0 | 35.48 | POS | POS |
| 5 | NEG | NEG | 0 | 0 | NEG | 0 | 0 | NEG | NEG | 1264 | POS | 33.3 | 35.7 | POS | 31.15 | 32.62 | POS | POS |
| 6 | NEG | NEG | 0 | 0 | NEG | 0 | 0 | NEG | NEG | 1300 | POS | 25.4 | 27.6 | POS | 27.31 | 27.55 | POS | POS |
| 7 | NEG | NEG | 0 | 0 | NEG | 0 | 0 | NEG | NEG | 1305 | POS | 23.8 | 26.2 | POS | 23.52 | 23.9 | POS | POS |
| 8 | NEG | NEG | 0 | 0 | NEG | 0 | 0 | NEG | NEG | 1306 | POS | 24.5 | 26.7 | POS | 24.83 | 25.09 | POS | POS |

Abbreviations: cycle threshold (Ct); relative light unit (RLU); nasopharyngeal (NP); oropharyngeal/bilateral nares (OPN).

## References

1. Bwire GM, Majigo MV, Njiro BJ, Mawazo A. Detection profile of SARS□CoV□2 using RT□PCR in different types of clinical specimens: A systematic review and meta□analysis. J Med Virol. 2021;93(2):719–725. doi:10.1002/jmv.26349

2. LeBlanc JJ, Heinstein C, MacDonald J, Pettipas J, Hatchette TF, Patriquin G. A combined oropharyngeal/nares swab is a suitable alternative to nasopharyngeal swabs for the detection of SARS-CoV-2. Journal of Clinical Virology. 2020;128:104442. doi:10.1016/j.jcv.2020.104442

3. Patriquin G, Davis I, Heinstein C, MacDonald J, Hatchette TF, LeBlanc JJ. Exploring alternative swabs for use in SARS-CoV-2 detection from the oropharynx and anterior nares. Journal of Virological Methods. 2020;285:113948. doi:10.1016/j.jviromet.2020.113948

4. World Health Organization. Tracking SARS-CoV-2 variants. Accessed January 16, 2022. https://www.who.int/emergencies/what-we-do/tracking-SARS-CoV-2-variants

5. Collie S, Champion J, Moultrie H, Bekker LG, Gray G. Effectiveness of BNT162b2 Vaccine against Omicron Variant in South Africa. N Engl J Med. Published online December 29, 2021:NEJMc2119270. doi:10.1056/NEJMc2119270

6. Araf Y, Akter F, Tang Y dong, Fatemi R, Parvez MdSA, Hossain Md G. Omicron variant of SARS-CoV-2: Genomics, transmissibility,and responses to current COVID-19 vaccines. Journal of Medical Virology. 2022;Epub ahead of print. doi:10.1002/jmv.27588

7. CDC COVID-19 Response Team. SARS-CoV-2 B.1.1.529 (Omicron) Variant — United States, December 1–8, 2021. MMWR Morb Mortal Wkly Rep. 2021;70(50):1731–1734. doi:10.15585/mmwr.mm7050e1

8. Huang N, Pérez P, Kato T, et al. SARS-CoV-2 infection of the oral cavity and saliva. Nat Med. 2021;27(5):892–903. doi:10.1038/s41591-021-01296-8

9. Marais G, Hsiao N yuan, Iranzadeh A, et al. Saliva Swabs Are the Preferred Sample for Omicron Detection. MedRxiv Infectious Diseases (except HIV/AIDS); 2021. doi:10.1101/2021.12.22.21268246

10. Iacobucci G. Covid-19: Runny nose, headache, and fatigue are commonest symptoms of omicron, early data show. BMJ. Published online December 16, 2021:n3103. doi:10.1136/bmj.n3103

11. Adamson B, Sikka R, Wyllie AL, Premsrirut P. Discordant SARS-CoV-2 PCR and Rapid Antigen Test Results When Infectious: A December 2021 Occupational Case Series. Infectious Diseases (except HIV/AIDS); 2022. doi:10.1101/2022.01.04.22268770

12. Goodall BL, LeBlanc JJ, Hatchette TF, Barrett L, Patriquin G. Investigating Sensitivity of Nasal or Throat (ISNOT): A Combination of Both Swabs Increases Sensitivity of SARS-CoV-2 Rapid Antigen Tests. MedRxiv Infectious Diseases (except HIV/AIDS); 2022. doi:10.1101/2022.01.18.22269426

13. LeBlanc JJ, Pettipas J, Di Quinzio M, Hatchette TF, Patriquin G. Reliable detection of SARS-CoV-2 with patient-collected swabs and saline gargles: A three-headed comparison on multiple molecular platforms. Journal of Virological Methods. 2021;295:114184. doi:10.1016/j.jviromet.2021.114184

